# Increased structural lung changes in adults with a history of bronchopulmonary dysplasia

**DOI:** 10.64898/2026.05.01.26352222

**Authors:** Melvin Pourbazargan, Kristina Piontkovskaya, Petra Um-Bergström, Anders Svensson-Marcial, Anders Lindén, Rebecka Stern, Eva Berggren Broström, Erik Melén, Iryna Kolosenko, Åsa M. Wheelock, Reza Karimi, Sven Nyren, C Magnus Sköld

**Author notes:** **Lung Obstruction in Adulthood of Prematurely Born (LUNAPRE) study**: https://clinicaltrials.gov/study/NCT02923648. Corresponding author: Kristina Piontkovskaya, Address: Lung and Allergy Clinic at Karolinska University Hospital, Solna, Gaevlegatan 55, Stockholm, 17164.

## Abstract

Bronchopulmonary dysplasia (BPD) in infancy is a risk factor for obstructive lung disease in adults. We hypothesized that adults born preterm and diagnosed with BPD have an altered lung architecture which is correlated to lung function impairment.

Individuals from the LUNAPRE cohort (clinicaltrials.gov/ct2/show/NCT02923648) were included: preterm born (gestational week <32) with (n=24) or without (n=23) a previous diagnosis of BPD, full term born with allergic asthma (n=22) and healthy volunteers (n=24). Inspiratory and expiratory HRCT scans were performed and interpreted by two expert reviewers in a blinded manner. Structural changes were scored and quantitative density measurements were analysed automatically using a dedicated post-processing workstation.

The HRCT scores were significantly higher in the BPD group compared to the other groups (p<0.001) and had highest numbers in subjects diagnosed with severe BPD. Most common HRCT changes were small peripheral opacities. Hypoattenuation during inspiration was only observed in the BPD group. Architectural distortion was observed in 6/24 BPD and 2/23 premature without BPD. HRCT scores correlated to FEV_1_ in a negative manner for preterm (p<0.001) and BPD (p<0.05) groups. Oxygen supplementation during the neonatal period correlated with HRCT score in a positive manner for preterm group (p<0.001). No differences in lung density were observed between the groups.

Young adults previously diagnosed with BPD have structural changes on CT which correlate with airway obstruction. Severity of BPD at the diagnosis was associated with CT abnormalities in adulthood. HRCT changes in adults with BPD were correlated with spirometry findings.

## INTRODUCTION

In 1967, Northway et al demonstrated changes in chest x-ray of preterm new-borns exposed to mechanical ventilation and highly concentrated oxygen treatment(1). They described clinical, radiological and pathological signs of a disease called hyaline-membrane disease and further adopted the name of bronchopulmonary dysplasia (BPD)(1). Since then, advancement in neonatal care have led to significantly improved care of children with BPD mainly by shortening the time of ventilation, less oxygen, use of antenatal corticosteroids and administration of surfactant. This has led to considerable improvement in the outcome of preterm-born infants with BPD(2, 3).

In earlier years paradigm of mechanical injury of alveoli due to mechanical ventilation in BPD has been shifted to formation of changes due to global alveolar arrest, a new form of BPD.

The knowledge of lung, airways and related vessels maturation in extreme preterm is limited. Severe chronic respiratory diseases and vascular diseases may arise from a preterm lung or severe BPD -due to the limitations in current knowledge, we do not know how the lung of a BPD child is going to look inside and out, so to say, referring mostly to the radiological consequences as well as spirometry changes (4). It is presumed that BPD lung is developing obstructive changes thorough the life. Most probably, leading to shortening of life and lowering quality of life.

Radiological and spirometry estimation of lung changes is crucial in an adult BPD lung for understanding of BPD/preterm lung physiology.

BPD is the most common cause of lung disease in preterm new-borns and is considered a chronic condition associated with airway obstruction that persists during adolescence, young adulthood and even later in life (5–9). Available evidence suggests that these individuals are predisposed to an early onset of COPD (10).

Lung maturation disruption is multifactorial and involves maternal, placental, fetal, and postnatal factors. Therefore, mechanical theory of BPD does not suffice anymore. Premaure lung involves as well changes in vessels in alveoli (11). Due to the complexity of BPD pathogenesis, we hypothesized that adult subjects born prematurely have an altered lung architecture as well as individuals with BPD. To address this hypothesis, we characterized qualitative and quantitative changes in the lungs of young adults born prematurely with or without BPD, utilizing high-resolution computed tomography (HRCT). We compared our findings with corresponding data from control groups - term-born healthy never-smokers and from subjects with mild allergic asthma.

## MATERIALS AND METHODS

### Study Subjects

A total of 93 individuals in four groups from the LUNg Adult PREmaturity (LUNAPRE) cohort (clinicaltrials.gov/ct2/show/NCT02923648) were included in the study (Table 1): Individuals born preterm (<32 gestational weeks) with (n=24) or without (n=23) a previous diagnosis of BPD, mild allergic asthma (>37 gestational weeks, n=22) and healthy controls (>37 gestational weeks, n=24). BPD was defined according to *Jobe and Bancalari* by the need for oxygen at 28 days of gestation and further classified as mild (grade 1), moderate (grade 2) or severe (grade 3) according to the need for oxygen at 36 weeks of gestation(12). According to this classification 9 subjects were diagnosed with mild BPD; 7 subjects with moderate BPD and 8 subjects with severe BPD. HRCT was performed at a median age of 19.8 years. The original LUNAPRE cohort included 96 individuals; 26 subjects with BPD, 23 subjects born preterm without BPD, 23 individuals with asthma and 24 healthy control individuals (13). Two subjects with BPD and one subject with asthma did not perform HRCT, but there were no significant differences in subject characteristics or lung function measurements observed in this study population. Demographic- and lung function characteristics are presented in an earlier paper (13) and are also briefly described in Table 1. The median age of the participants at the time of HRCT was 19.8 years, the two preterm groups were slightly younger at the time of HRCT examination. The sex distribution was very similar across the groups. The BPD group had lower FEV_1_, and FEV_1_/FVC ratio compared to the other groups. Lung clearance index (LCI) was decreased in BPD group (BPD vs healthy p<0.01). Perinatal characteristics are presented in supplemental Table E1. Subjects with BPD had lower gestational age (median 26 weeks (24–31) vs 29.5 weeks (27–32)) and birth weight (median 951g (580-2136) vs 1460 g (710-2200) compared to the preterm group.

**Table 1.**
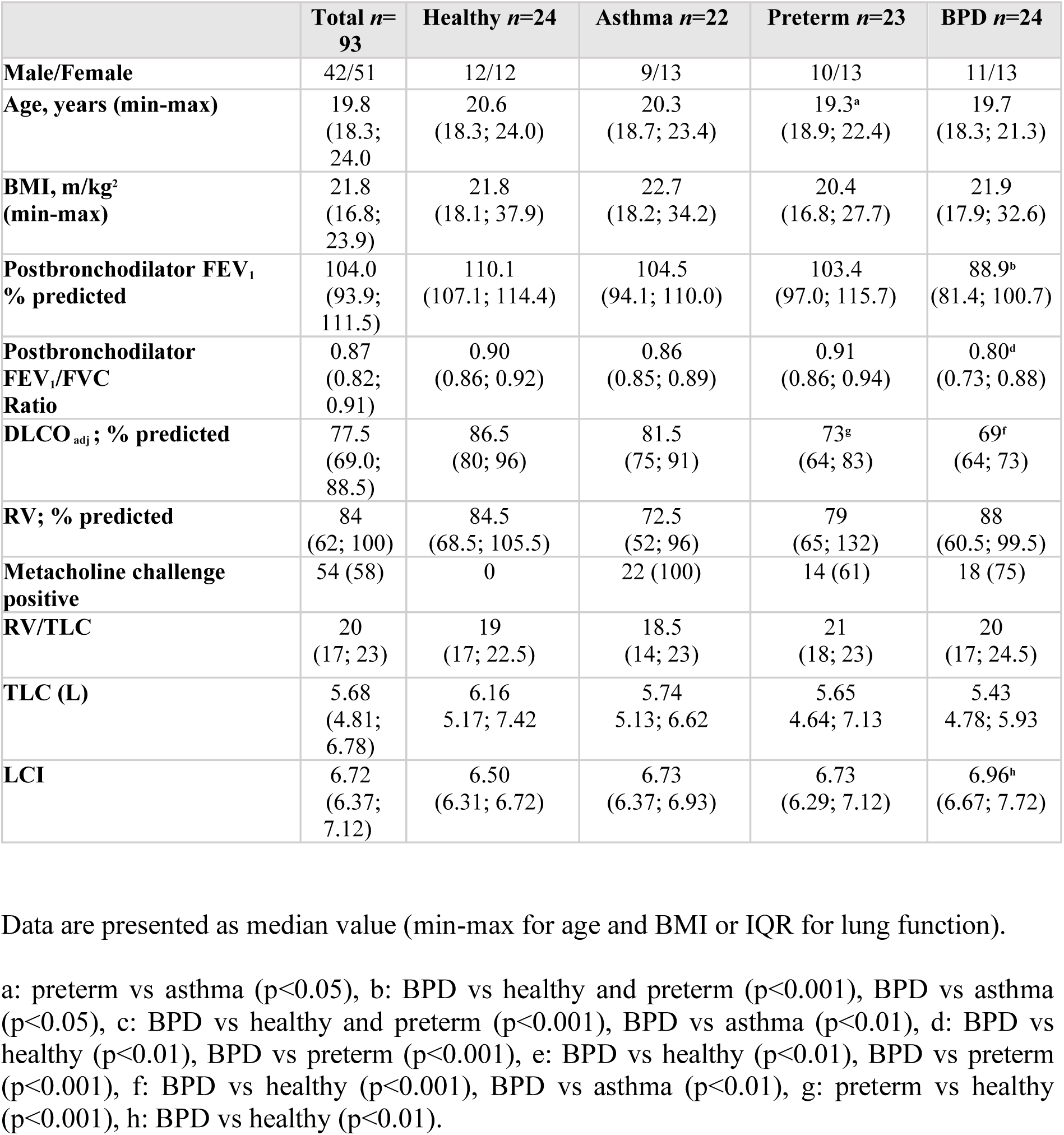
Demographics and lung function of study participants.

All participants provided written informed consent, and the study was approved by the Swedish Ethical Review Authority (ref: 2012/11872-31/4, ref: 2013/1416-32, ref 2017/868/-32) and from the Swedish Radiation Protection Committee (ref K2641-2012).

### High-resolution computed tomography (HRCT)

93 participants underwent HRCT, 85 by Simens Somatom Sensation 16 slice CT (Siemens Healthcare, Forcheim, Germany) at the department of Radiology, Karolinska University Hospital, Solna, Sweden. Due to a change of equipment during the inclusion period, 8 participants underwent HRCT using Simens Somatom Definition Flash 128 slice CT (Siemens Healthcare, Forchheim, Germany) at the department of Pediatric Radiology, Childrens Hospital, Karolinska University Hospital Solna, Sweden. Scanning parameters were modified to make images from different scanners as similar as possible. CT scans were performed fully inhaled and 6 single scans were taken during forced expiration at 3 levels (4 cm above carina, at the level of carina and 4 cm below carina) A low dose protocol was used to comply with the restrictions of a radiation dose to < 1 mSv. For both scanners 120 kV was used, reconstructed with kernel B30, B40 and B70 in 1 mm thick slices with 0,5 mm increment. Radiation doses were kept down by individually reducing mA. The two CT scanners differed with Siemens Somatom Sensation having 16 x 0,75 mm and Definition Flash having 128 x 0,5 mm collimation.

### Qualitative HRCT analysis

On the basis of results from previous studies on children and young adults with BPD(5, 14, 15), we used a modified scoring system including 12 features (supplementary Table E2). CT scans were then analyzed at lobar level where the lingula was considered a separate lobe. Linear-, triangular-, reticular subpleural lung opacities, local hypoattenuation on inspiration/mosaic pattern, emphysema, architectural distortion, bronchiectasis, consolidation, parenchymal noduli and subpleural noduli were scored from 1 to 6 points based on the number of affected lobes. Each of the above-mentioned findings could thereby generate 1 point for each lobe, regardless of if multiple findings were found. Air trapping and bronchial wall thickening were scored as 0 (absent) and 1 (present). The score range was thereby 0 – 62 points for each study participant for all the findings in the lungs. Abnormal lung features are defined according to the nomenclature in Fleischner Society: Glossary of Terms for thoracic Imaging (16) (supplementary Table E2).

CT scans were primarily evaluated blinded by two expert examiners, one radiologist (SN) and one pulmonologist (RK), both with >10 years’ experience in rating thoracic CT-scans. The total number of potential lung abnormalities each examiner assessed was 5766. After primary evaluation, the examiners together re-evaluated 235 findings that were assessed in disagreement for conclusion. The HRCT scoring system yielded an interobserver ratio with a weighted kappa value of 0.32 (CI; 0,26-0,39), a positive predictive value of 98% and the sensitivity to 98%.

### Quantitative HRCT analysis

Quantitative measurements of the CT images on 85 participants were made using a dedicated post-processing workstation (Syngo.Via CT Pulmo 3D, VB50, Siemens Healthcare). This program measures total lung volume, mean Hounsfield unit (HU) and percent of lower attenuation value (LAV%) less than 950 HU. Lung regions with an attenuation <-950 HU were regarded as emphysema (17). Due to different CT modalities used for 8 participants that underwent scans on another CT (see above), quantitative measurements could not be assessed on these individuals. From the 85 subjects, data could not be extracted from 1 healthy participant and the LAV% could not be extracted from 1 asthmatic and 2 preterm subjects.

### Statistical methods

Continuous variables were summarized as median (interquartile range [IQR]). For non-normal variables, group comparisons were made using a non-parametric analysis of variance (Kruskal Wallis test) using Dunn’s correction for multiplicity. Categorical variables were summarized as proportions. Linear regression modeling (Spearman) was used for correlation measurements. The IBM SPSS Statistics version 27 (IBM, Armonk, NY) and GraphPad Prism version 8.2.0 (San Diego, CA, USA) were used. A p-value ≤0.05 was considered as statistically significant.

## RESULTS

### Qualitative HRCT findings

The distribution of all observed findings is listed in Table 2, and the total HRCT score is shown in Figure 1. The BPD group displayed more HRCT findings compared with the other groups (p<0.001 for all comparisons (Figure 1). Most of the observed changes were small peripheral opacities defined as sub-pleural triangular and linear opacities (Figure 2 a-b). Even though more common in the BPD group (71% of individuals for linear and 54% for triangular opacities, p<0.01) these findings were also found in in healthy controls (29% linear, 13% triangular), asthma (18% linear, 18% triangular) and premature individuals without BPD (9% linear, 26% triangular) (Table 2).

**Figure 1.**
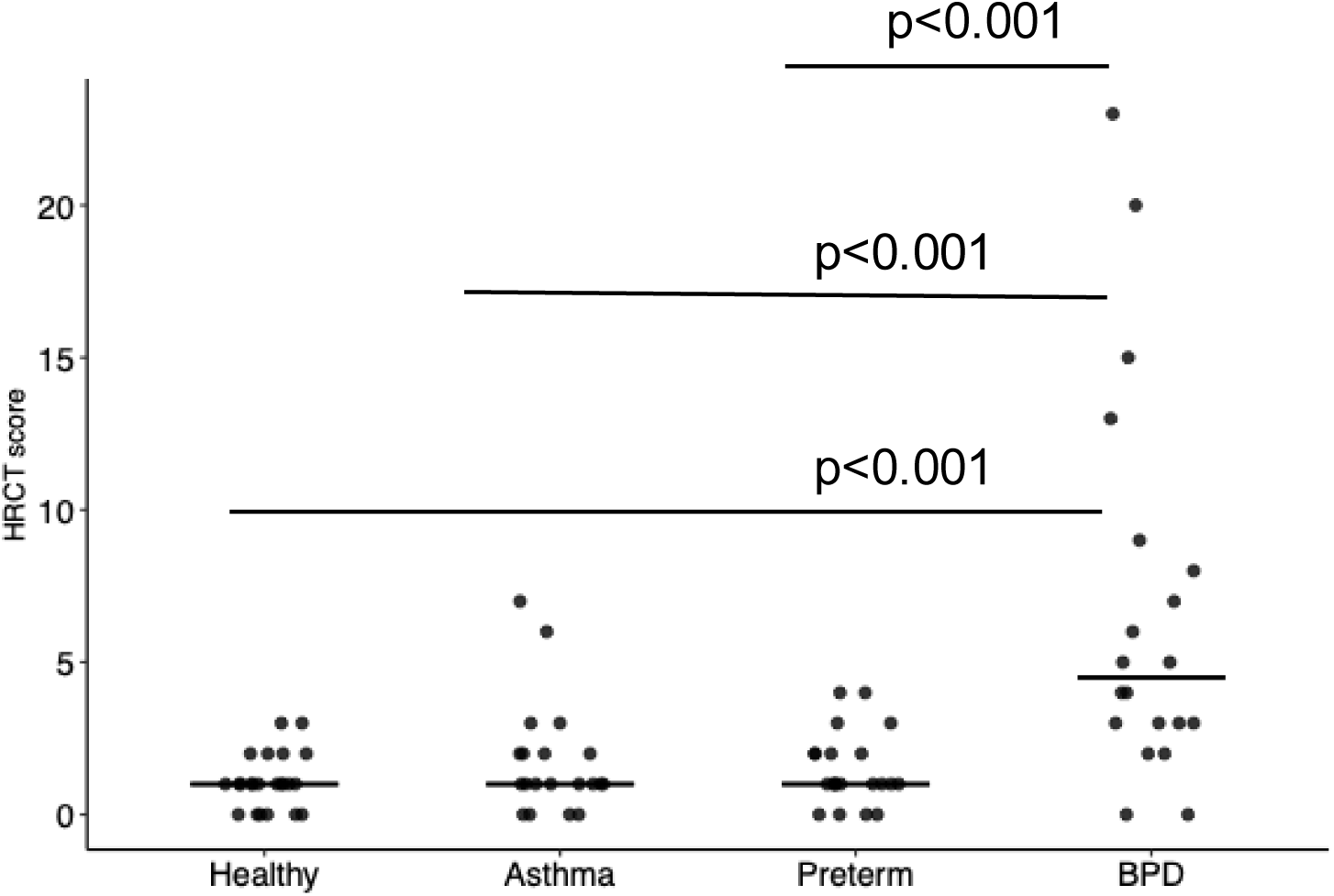
Total HRCT score for healthy, asthma, preterm (non-BPD) and BPD subjects. BPD are subdivided into grade 1 (•), grade 2 (□), and grade 3 (♦), see text. Non-parametric analysis of variance (Kruskal-Wallis test) using Dunns correction for multiplicity.

**Figure 2.**
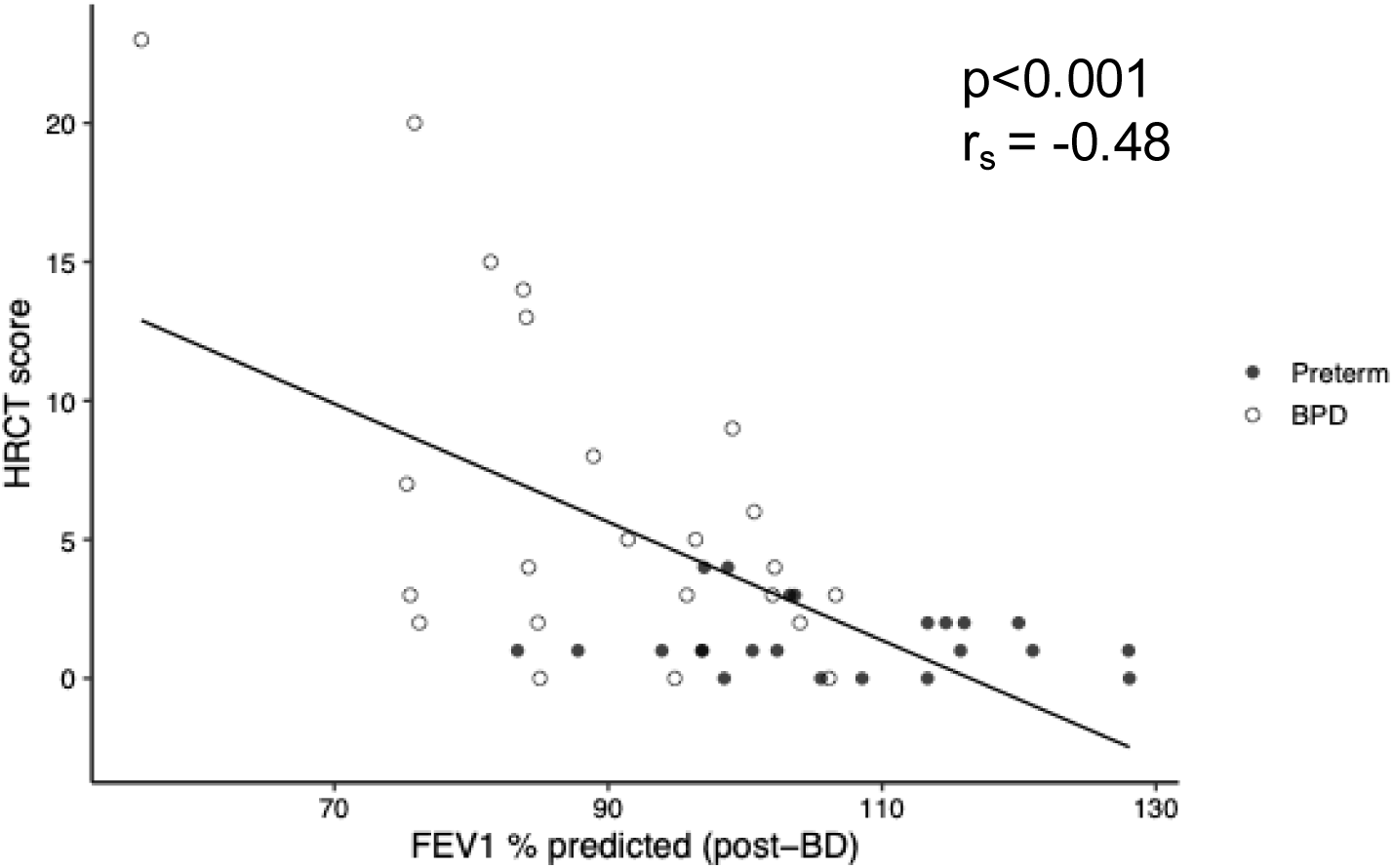
a-g. Typical structural changes observed on HRCT in patients diagnosed with BPD. a) Subpleural triangular lung opacities, b) Subpleural linear opacity, c) Local hypoattenuation on inspiration d) air trapping, e) bronchial wall thickening, f) architectural distortion (white arrow), local hypoattenuation (star), subpleural triangular opacities (black arrow), g) subpleural noduli.

**Table 2.**
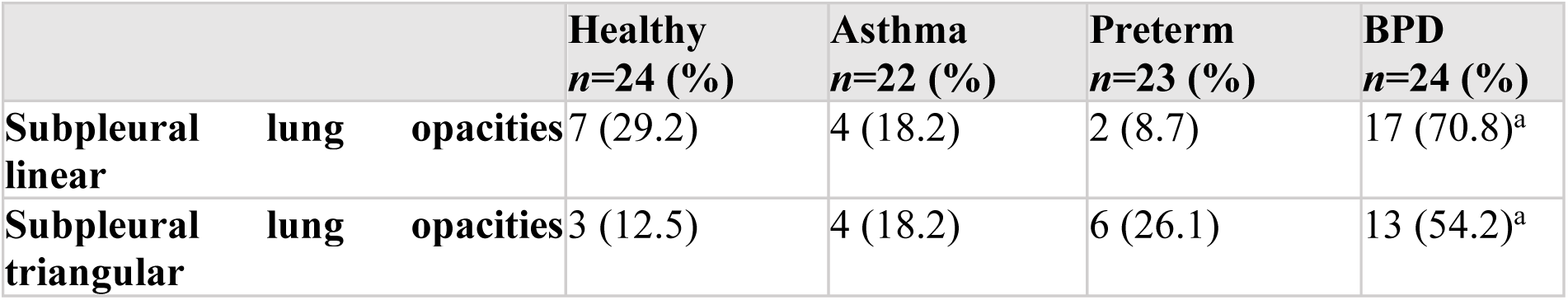
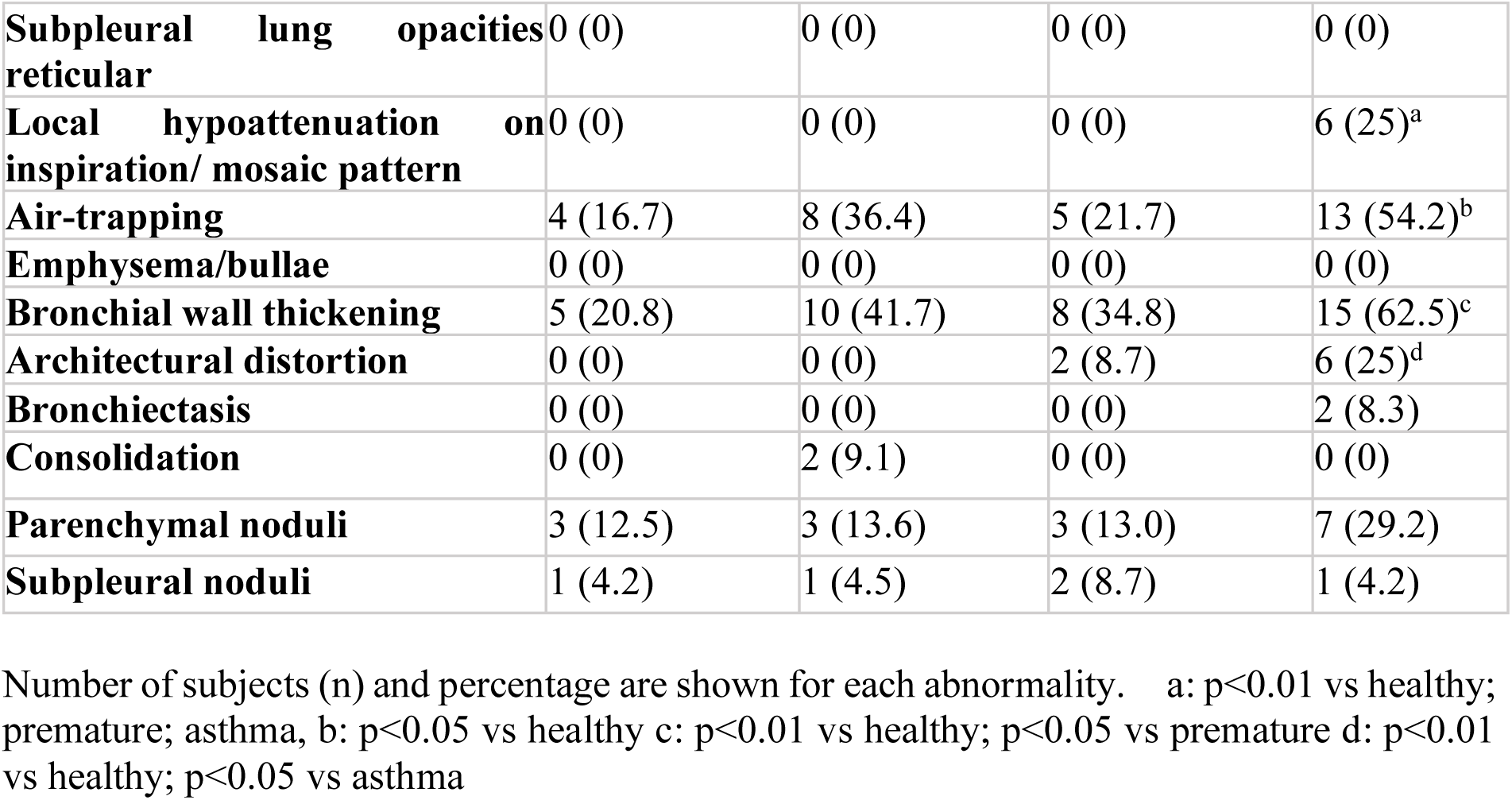
Quantitative HRCT findings in healthy, asthma, preterm and BPD.

Hypoattenuation during inspiration, also referred to as mosaic perfusion, was distinguished from air trapping and evaluated in end-inspiratory scans (Figure 2c). Hypoattenuation was observed in the BPD group only, in six subjects (25% of the BPD group, p<0.05).

Air trapping (54%) (Figure 2d) and bronchial wall thickening (63%) (Figure 2e) were more common in the BPD group (p<0.05) compared to healthy controls (17% and 21%) and premature without BPD (22% and 35%). Architectural distortion (Figure 2f) was observed in 6 subjects in the BPD group (25%, p<0.05 compared to healthy and individuals with asthma) and in two subjects in the preterm group. Subpleural noduli (Figure 2g) were represented in all groups without any significant difference between the groups.

Subjects with BPD grade 3 demonstrated increased HRCT scores (Figure 1), (median score 9 SD: 8.3 compared to BPD grade 2 (median score 6 SD:4.9) and BPD grade 1 (median score 3 SD: 2.1) In the six cases of local hypoattenuation, five subjects had BPD grade 3 and one subject had BPD grade 2. BPD subjects with hypoattenuation showed significantly lower FEV1 % predicted compared to subjects without hypoattenuation (p<0.01). In six cases of architectural distortion in the BPD group, three subjects were diagnosed with BPD grade 3, one subject with BPD grade 2 and two subjects with BPD grade 1. Subjects with BPD grade 2 and 3 with architectural distortion also showed findings of local hypoattenuation on inspiration.

### Quantitative HRCT findings

There were no differences in automatic CT lung volume measurements between the groups. BPD subjects showed numerically lower lung volumes compared to healthy subjects (median value 5240 mL vs 6371 mL (Table 3), but this difference was not statistically significant. The total lung volume measured with HRCT correlated with the spirometry measurements of TLC (r_s_ 0,91 p<0.001). The lung attenuation values were similar for the BPD ( -870 HU), healthy subjects (-879 HU), asthma (-874 HU) and preterm (-879 HU). Percentage of the lung with a low attenuation value <950 HU (LAV%) did not show any substantial group differences (Table 3).

**Table 3.** Lung volume, mean attenuation and LAV% in healthy, asthma, preterm and BPD.

|  | **Total n=84** | **Healthy n=23** | **Asthma n=20** | **Preterm n=21** | **BPD n=20** |
| --- | --- | --- | --- | --- | --- |
| **CT volume total, mL** | 5407<br>(4635; 6727) | 6371<br>(5032; 7295) | 5572<br>(4392; 6531) | 5167<br>(4485; 7022) | 5240<br>(4808; 5874) |
| **Attenuation, HU (mean)** | -877<br>(-887; -865) | -874<br>(-885; -870) | -874<br>(-883; -852) | -879<br>(-892; -862) | -879<br>(-890; -862) |
|  | **Total n=81** | **Healthy n=23** | **Asthma n=19** | **Preterm n=19** | **BPD n=20** |
| **LAV (%)** | 19.0<br>(14.0; 24.2) | 18.3<br>(14.8; 24.2) | 19.6<br>(12.9; 23.8) | 19.0<br>(14.6; 25.0) | 22.4<br>(13.6; 28.3) |

HU=mean Hounsfield attenuation of the lungs; CT=computed tomography; LAV%: percent of lung with low attenuation value ≤ 950 HU
Data are presented as median (interquartile range IQR)

### Correlations between quantitative and qualitative HRCT measurements, neonatal data and lung function

The qualitative HRCT score negatively correlated with FEV_1_ for all preterm subjects (p<0.001, r_s_-0.48) (Figure 3) and for BPD subjects (p<0.05, r_s_-0.46).

**Figure 3.**
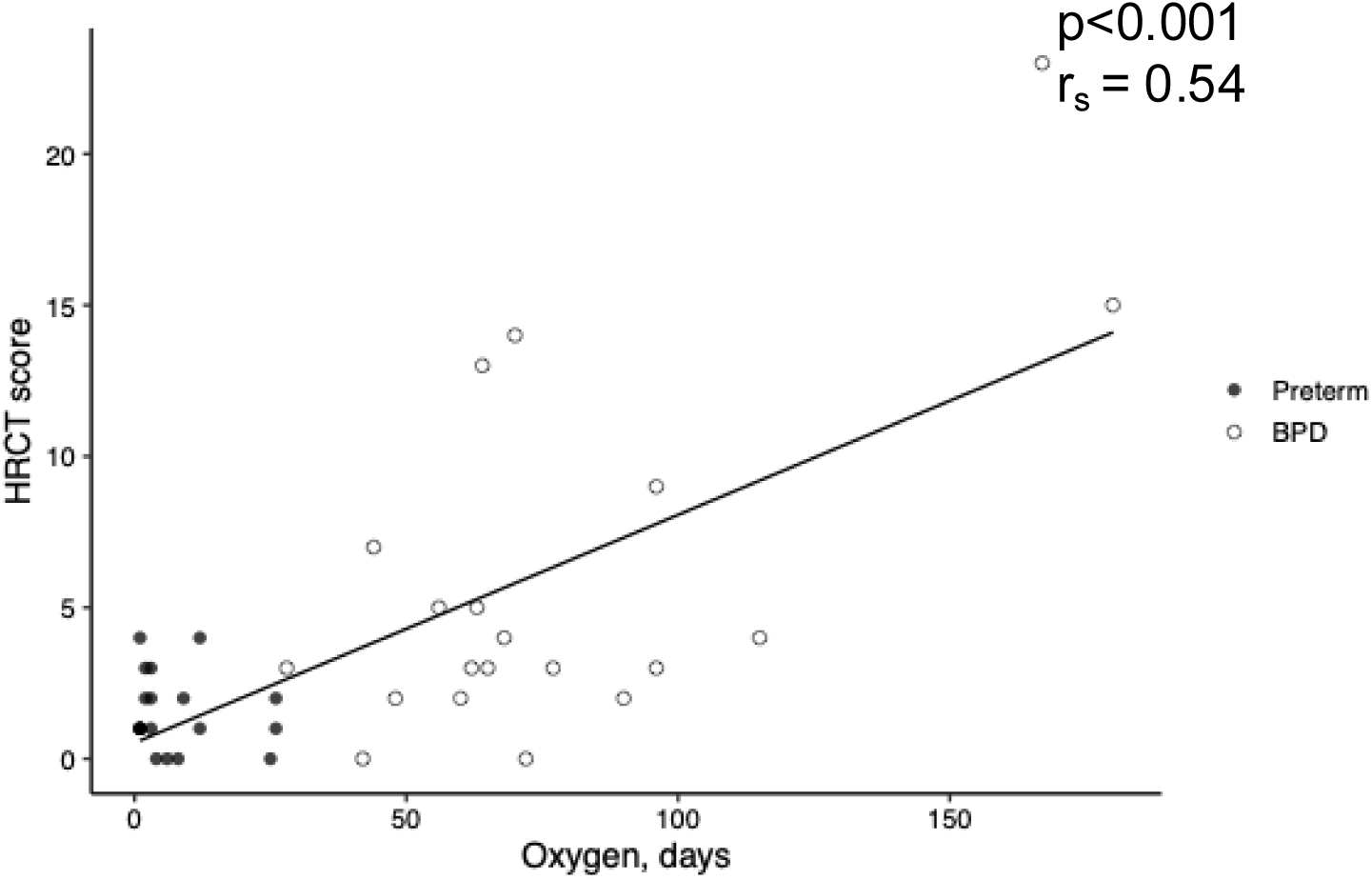
Correlation between HRCT score and FEV_1_ % predicted for all preterm subjects. Full black circle represents preterm w/o BPD (•), White circle represents BPD (□). Analysis: Linear regression modeling (Spearman) for correlation measurements (r_s_).

Oxygen delivery during the neonatal period was correlated with HRCT score for both preterm and BPD subjects (p<0.001 r_s_ 0.54) (Figure 4) and BPD alone (p<0.095 r_s_ 0.38). In four persons with BPD, data were missing regarding oxygen treatment during the neonatal period, in one subject data on invasive ventilation was missing, and in one subject - days on non-invasive ventilation. Both noninvasive (CPAP) and invasive ventilation during the neonatal period were correlated with both the preterm groups to HRCT score (p<0.05 r_s_ 0.35) (Figure 5).

**Figure 4.**
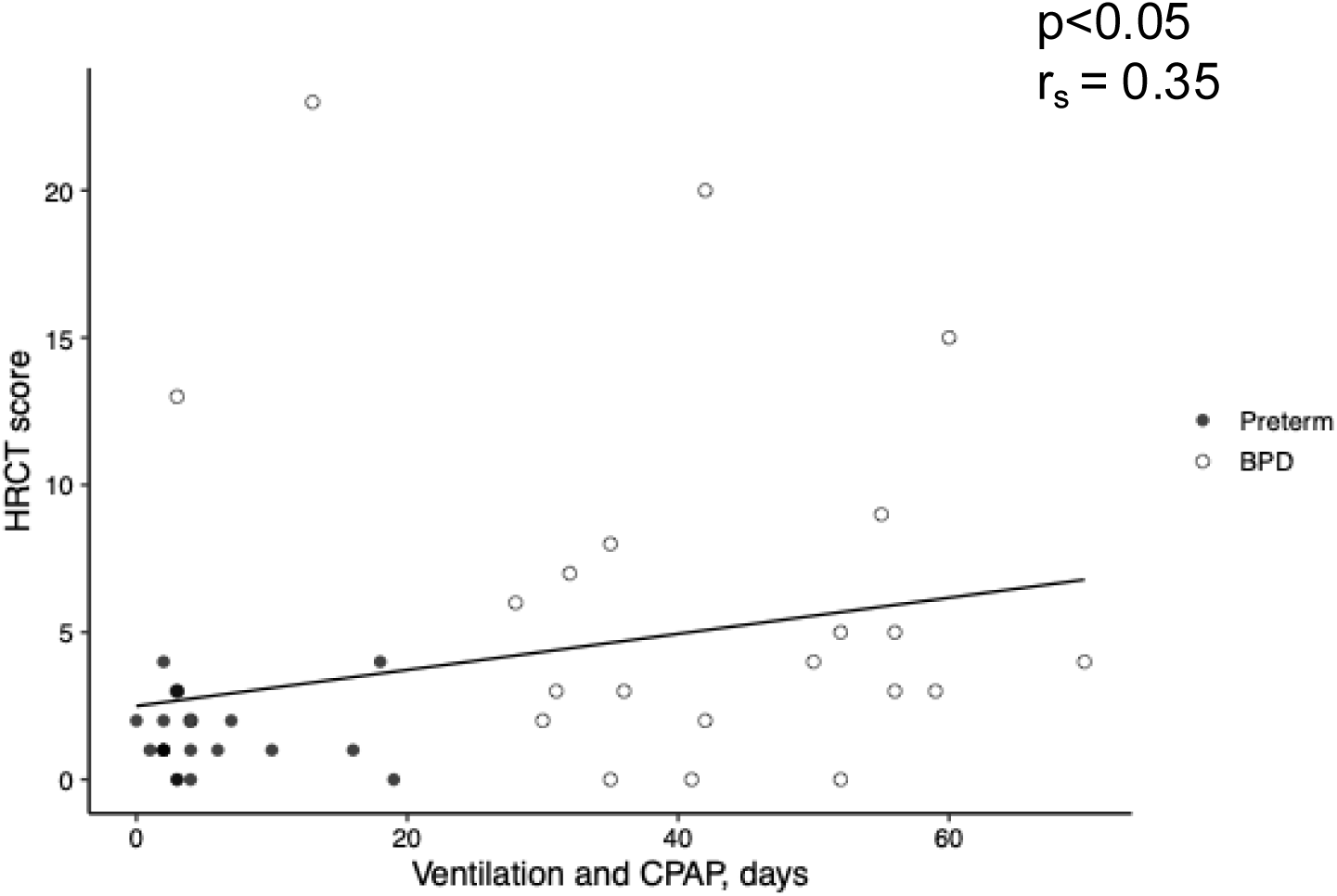
Correlation between HRCT score and days on oxygen during the neonatal period in preterm born subjects. Full black circle represents preterm w/o BPD (•), White circle represents BPD (□). Linear regression modeling (Spearman) for correlation measurements (r_s_).

**Figure 5.**
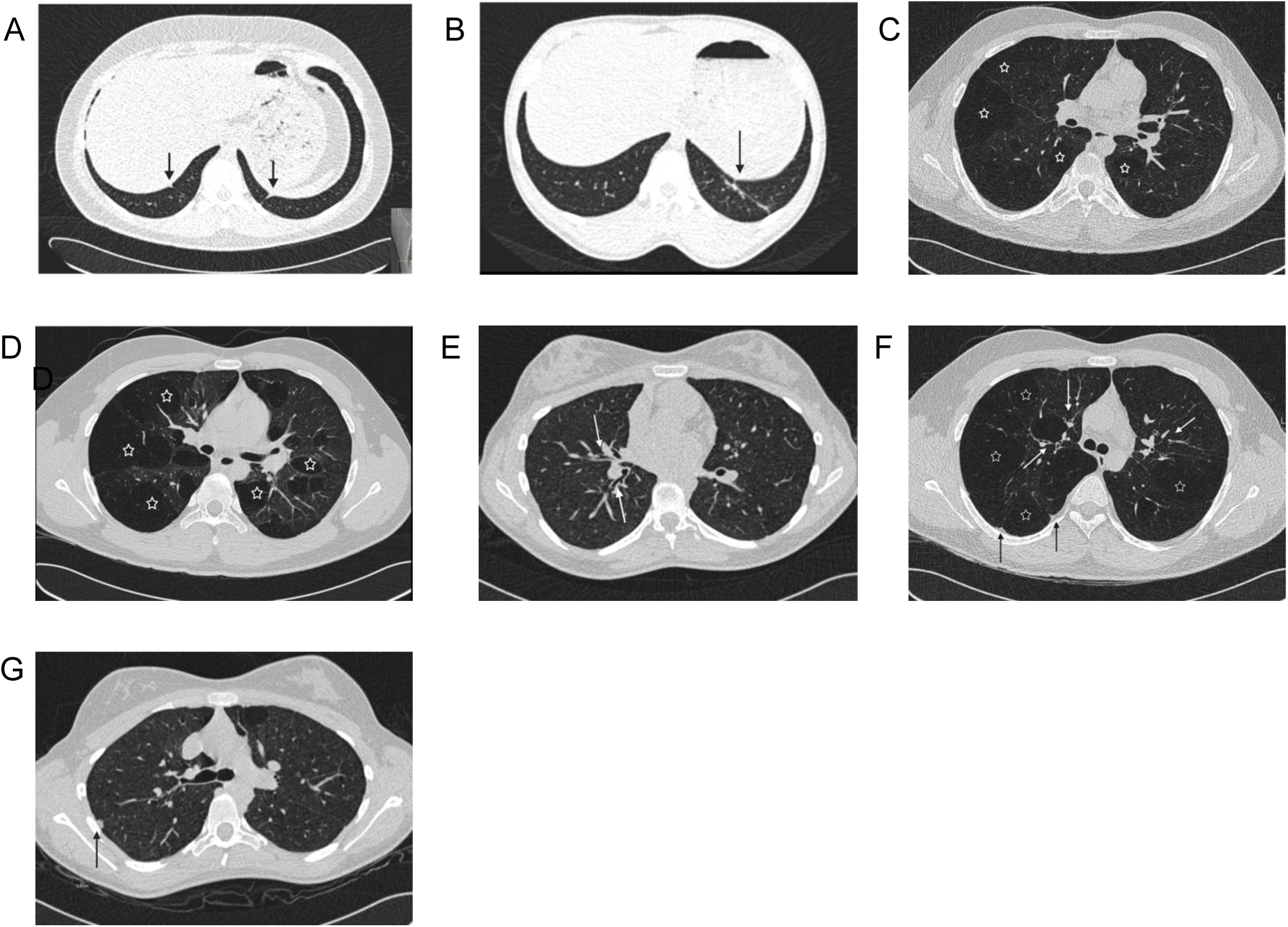
Correlation between HRCT score and days of noninvasive (CPAP) and invasive ventilation (sum of days on CPAP and invasive ventilation). Subjects include both BPD and preterm subjects. Full black circle represents preterm w/o BPD (•), White circle represents BPD (□). Linear regression modeling (Spearman) for correlation measurements (r_s_).

HRCT score was negatively correlated to diffusion capacity for all subjects (p<0.001, r_s_ 0,37) (supplemental figure E4).

There was no statistically significant correlation between CT lung volumes and HRCT scores (r_s_ 0,004 p=0,97). However, a moderate correlation between CT lung volumes and diffusion capacity (DLCOadj) was observed (r^s^ 0,46 p=0,001) (supplemental Figure E1).

There were no statistically significant correlations between density measurements (mean HU and LAV%) and diffusion capacity in the total study group (r_s_ 0,06, p=0,60 for mean HU and r_s_ 0,60, p=0,95 for LAV%) the pooled preterm group (r_s_ 0,099, p=0,56) or in the BPD group alone (r_s_ 0,34, p=0,16 for mean HU and r_s_ -0,35, p=0,14 for LAV%) (supplemental Figures E2 and E3).

## DISCUSSION

According to our data young adults previously diagnosed with BPD in the neonatal period had more structural changes in HRCT compared to prematurely born adults without BPD, asthma and healthy controls. The most common were local hypoattenuation, subpleural opacities, bronchial wall thickening and air-trapping. The structural findings correlated negatively with lung function and positively with days on oxygen during the neonatal period.

Previous studies have shown that young adults with BPD have an obstructive lung function impairment and low DLCO later in life(13, 18–20). The first study of CT scans on children diagnosed with BPD was done by Oppenheim *et al* already in 1994(21). They studied 23 subjects with a mean age of 4 years and described areas of hypoattenuation and well-defined linear and triangular sub-pleural densities. Another study (11) on children aged 6-8 years showed persistent structural abnormalities in chest CT. Aukland *et al* performed chest CT scans comparing new BPD (mean age 10 years) and old BPD (mean age 18 years) and found no differences between the two BPD groups in the HRCT score(22).

Mastrigt *et al* (19) presented a literature review to give an overview of the scoring system used to describe structural lung abnormalities in subjects with BPD. The study concluded that the most sensitive structural abnormality associated with BPD severity is hypoattenuation on inspiration.

We believe that the structural findings described in the present and previous studies may be signs of abnormalities in the distal airways resulting in reduced elastic recoil leading to airflow limitation. Architectural distortion, reflecting a disturbed appearance of the lung due to abnormal displacement of pulmonary structures, was only found in the preterm groups. The clinical consequences of this finding are unknown, but it shows a clear disarrangement of the normal lung. Two subjects born preterm without BPD and six subjects with BPD showed architectural distortion affecting more than two lobes. These individuals had higher total CT score compared to the mean score for all BPD subjects. The same pattern is seen in subjects with local hypoattenuation on inspiration affecting more than two lobes. Subjects with local hypoattenuation and architectural distortion affecting more than one lobe also show significantly lower lung function when measuring FEV1 compared to BPD subjects without these findings. This apparent abnormality in the lung parenchyma and maldistribution of ventilation seems to be more common to the more severe form of BPD and a risk factor for obstructive lung function impairment.

We assumed that the decreased diffusion capacity in the BPD subjects could be reflected by early-stage emphysema(23, 24). However, emphysema was not present neither in the quantitative scoring or in the qualitative measurement of LAV%. It has previously been shown that LAV% is a poor indicator of mild emphysema(25). We did not find any significant difference between the groups when measuring the total lung volume by CT, but the preterm groups showed lower mean volumes compared to the healthy group and individuals with asthma. Studies have suggested that BPD individuals are subjects to alveolarisation arrest in the neonatal period(26, 27), leading us to the hypothesis that this could be reflected by smaller lungs leading to decreased diffusion capacity in these subject. Looking at the correlation between lung volume and DLCOadj, a strong correlation is evident with low CT volume and lower diffusing capacity.

With access to perinatal data, we tried to identify correlations to events in an early stage of life reflecting the degree of abnormal CT findings leading to airflow limitation. Santema *et al* (28) compared BPD to a non-BPD group and lung function measurements including chest CT density measurements in young adult age, but were not able to pinpoint a perinatal characteristic that could predict lower lung function in adulthood. We found a significant correlation between days on oxygen and ventilator support and HRCT score.

The strengths of this study include the longitudinal comparison of perinatal data and data from young adult life including extensive examinations and data measurements of the LUNAPRE cohort. To our knowledge, this is the first study to evaluate the respiratory function, quantitative and qualitative HRCT measurements in BPD subjects compared to healthy controls, individuals with asthma and preterm born without BPD – enabling us to distinguish specific characteristics of BPD individuals.

Ideally this study would need a group of age matched smoking related COPD subjects for direct correlation of the findings. However, the ability to identify and recruit subjects with COPD related to smoking with the mean age of 19 years was beyond the scope of this study. Another limitation is the relatively small number of subjects reducing the power in our study.

Subjects diagnosed with BPD often have chronic respiratory disorders and lung function trajectories corresponding to COPD. In recent guidelines from the European Respiratory Society it is recommended that patients with BPD should be referred from a paediatric pulmonologist to an adult pulmonologist for long term care(4). There is a need to identify those individuals born premature who are of greatest risk to develop lung disfunction in their young adult life(29). Here, we have data suggesting that young adults diagnosed with BPD have increased structural lung abnormalities and that these findings correlate with decreased lung function. Those most affected are young adults diagnosed with severe BPD with prolonged need of ventilator support and need of supplemental oxygen treatment at birth. Further research in the adult BPD group may help us understand the long-term effects and to evaluate if these findings are persistent or rather progressive and hopefully further identify treatment possibilities for these patients.

## Data Availability

All data produced in the present study are available upon reasonable request to the authors

## Abbreviations list

BMI: body mass index
BPD: Bronchopulmonary dysplasia
DLCOadj: diffusion capacity of the lung for carbon monoxide adjusted for blood hemoglobin value percent predicted
F: female
FEV1: Forced expiratory volume in 1 second
FVC: forced vital capacity
HRCT score: high-resolution computer tomography score
HU: Hounsfield Units
IQR: interquartile range
LAV%: lower attenuation value
LCI: lung clearance index
LUNAPRE study: Lung Obstruction in Adulthood of Prematurely Born study
M: male
RV: residual volume percent predicted
TLC: total lung capacity Liter

## ACKNOWLEDGEMENTS

Many thanks to all research nurses: Heléne Blomqvist, Margitha Dahl, Gunnel de Forest, Emma Sundström, all colleagues involved in the LUNAPRE project and all the study participants for making this study possible.

## References

1. Northway WH Jr RR, Porter DY. PMID: 5334613. Pulmonary disease following respirator therapy of hyaline-membrane disease. Bronchopulmonary dysplasia. N Engl J Med 1967 Feb;16;276(7):357–68.

2. Wilson AC. What does imaging the chest tell us about bronchopulmonary dysplasia? Paediatric Respiratory Reviews. 2010;11(3):158–61.

3. Stoll BJ HN, Bell et al. Trends in Care Practices, Morbidity, and Mortality of Extremely Preterm Neonates. JAMA 2015 Sep 8;314(10):1039–51. 2015.

4. Duijts L, van Meel ER, Moschino L, Baraldi E, Barnhoorn M, Bramer WM, et al. European Respiratory Society guideline on long-term management of children with bronchopulmonary dysplasia. Eur Respir J. 2020;55(1).

5. Howling. Howling SJ, Northway WH Jr, Hansell DM, Moss RB, Ward S, Müller NL. Pulmonary sequelae of bronchopulmonary dysplasia survivors: high-resolution CT findings. AJR Am J Roentgenol. 2000 May;174(5):1323–6. doi: 10.2214/ajr.174.5.1741323. PMID: 10789786. 2000.

6. Broström EB, Thunqvist P, Adenfelt G, Borling E, Katz-Salamon M. Obstructive lung disease in children with mild to severe BPD. Respiratory Medicine. 2010;104(3):362–70.

7. Halvorsen T, Skadberg BT, Eide GE, Røksund OD, Carlsen KH, Bakke P. Pulmonary outcome in adolescents of extreme preterm birth: a regional cohort study. Acta paediatrica (Oslo, Norway : 1992). 2004;93(10):1294-300.

8. Baraldi E, Filippone M. Chronic lung disease after premature birth. The New England journal of medicine. 2007;357(19):1946–55.

9. Bui DS, Perret JL, Walters EH, Lodge CJ, Bowatte G, Hamilton GS, et al. Association between very to moderate preterm births, lung function deficits, and COPD at age 53 years: analysis of a prospective cohort study. Lancet Respir Med. 2022;10(5):478–84.

10. McGrath-Morrow SA, Collaco JM. Bronchopulmonary dysplasia: what are its links to COPD? Therapeutic Advances in Respiratory Disease. 2019;13:1753466619892492.

11. Durlak W, Thébaud B. The vascular phenotype of BPD: new basic science insights-new precision medicine approaches. Pediatr Res. 2024;96(5):1162–71.

12. Jobe AH BE. Bronchopulmonary dysplasia. Am J Respir Crit Care Med. 2001; Jun;163(7):1723–9.

13. Petra. Um-Bergström P, Hallberg J, Pourbazargan M, Berggren-Broström E, Ferrara G, Eriksson MJ, Nyrén S, Gao J, Lilja G, Lindén A, Wheelock ÅM, Melén E, Sköld CM. Pulmonary outcomes in adults with a history of Bronchopulmonary Dysplasia differ from patients with asthma. Respir Res. 2019 May 24;20(1):102. doi: 10.1186/s12931-019-1075-1. PMID: 31126291; PMCID: PMC6534852. 2019.

14. Aukland. Aukland SM, Halvorsen T, Fosse KR, Daltveit AK, Rosendahl K. High-resolution CT of the chest in children and young adults who were born prematurely: findings in a population-based study. AJR Am J Roentgenol. 2006 Oct;187(4):1012–8. doi: 10.2214/AJR.05.0383. PMID: 16985150. 2006.

15. Mastrigt. van Mastrigt E, Logie K, Ciet P, Reiss IK, Duijts L, Pijnenburg MW, Tiddens HA. Lung CT imaging in patients with bronchopulmonary dysplasia: A systematic review. Pediatr Pulmonol. 2016 Sep;51(9):975–86. doi: 10.1002/ppul.23446. Epub 2016 May 5. PMID: 27148803. 2016.

16. Hansell DM BA, MacMahon H, McLoud TC, Müller NL, Remy J. Fleischner Society: glossary of terms for thoracic imaging. Radiology 2008 Mar;246(3):697–722. 2008.

17. Lynch DA. Progress in Imaging COPD, 2004 - 2014. Chronic Obstr Pulm Dis. 2014;1(1):73–82.

18. Caskey S GA, Rowan S, Gillespie S, Clarke J, Riley M, Megarry J, Nicholls P, Patterson C, Halliday HL, Shields MD, McGarvey L. Structural and Functional Lung Impairment in Adult Survivors of Bronchopulmonary Dysplasia. . Ann Am Thorac Soc 2016. Aug;13(8):1262–70. doi: 10.1513/AnnalsATS.201509-578OC. PMID: 27222921.

19. Gough A, Linden M, Spence D, Patterson CC, Halliday HL, McGarvey LP. Impaired lung function and health status in adult survivors of bronchopulmonary dysplasia. Eur Respir J. 2014;43(3):808–16.

20. Vollsaeter M, Roksund OD, Eide GE, Markestad T, Halvorsen T. Lung function after preterm birth: development from mid-childhood to adulthood. Thorax. 2013;68(8):767–76.

21. Oppenheim C, Mamou-Mani T, Sayegh N, de Blic J, Scheinmann P, Lallemand D. Bronchopulmonary dysplasia: value of CT in identifying pulmonary sequelae. American Journal of Roentgenology. 1994;163(1):169–72.

22. Aukland SM, Rosendahl K, Owens CM, Fosse KR, Eide GE, Halvorsen T. Neonatal bronchopulmonary dysplasia predicts abnormal pulmonary HRCT scans in long-term survivors of extreme preterm birth. Thorax. 2009;64(5):405.

23. Pompe E, Galban CJ, Ross BD, Koenderman L, Ten Hacken NH, Postma DS, et al. Parametric response mapping on chest computed tomography associates with clinical and functional parameters in chronic obstructive pulmonary disease. Respir Med. 2017;123:48–55.

24. Bankier AA, De Maertelaer V, Keyzer C, Gevenois PA. Pulmonary emphysema: subjective visual grading versus objective quantification with macroscopic morphometry and thin-section CT densitometry. Radiology. 1999;211(3):851–8.

25. Vikgren J, Khalil M, Cederlund K, Sorensen K, Boijsen M, Brandberg J, et al. Visual and Quantitative Evaluation of Emphysema: A Case-Control Study of 1111 Participants in the Pilot Swedish CArdioPulmonary BioImage Study (SCAPIS). Acad Radiol. 2020;27(5):636–43.

26. Husain AN, Siddiqui NH, Stocker JT. Pathology of arrested acinar development in postsurfactant bronchopulmonary dysplasia. Hum Pathol. 1998;29(7):710–7.

27. Kinsella JP, Greenough A, Abman SH. Bronchopulmonary dysplasia. Lancet. 2006;367(9520):1421–31.

28. Santema HY, Stolk J, Los M, Stoel BC, Tsonaka R, Merth IT. Prediction of lung function and lung density of young adults who had bronchopulmonary dysplasia. ERJ Open Res. 2020;6(4).

29. Melén E, Faner R, Allinson JP, Bui D, Bush A, Custovic A, et al. Lung-function trajectories: relevance and implementation in clinical practice. Lancet. 2024;403(10435):1494–503.

