## Supplemental materials for "Increased structural lung changes in adults with a history of bronchopulmonary dysplasia"

**Tabel E1. Perinatal characteristics of the study subjects**

| | Healthy *n*=24 | Asthma *n*=22 | Preterm *n*=23 | BPD *n*=24 |
|---|---|---|---|---|
| **Male/Female** | 12/12 | 9/13 | 10/13 | 11/13 |
| **Gestational age at birth, weeks** | 40 (37-43 | 39.5 (38-42) | 29.5[b] (27-32) | 26[a] (24-31) |
| **Birth weight** | 3458 (2670-4550) | 3445 (2660-4840) | 1460[d] (710-2200) | 951[c] (580-2136) |
| **Maternal smoking during preagnancy** | 5 (21) | 2 (9) | 3 (13) | 5 (21) |
| **Prenatal corticosteroid therapy** | N/A | N/A | 12 (52) | 10 (42) |
| **Cesarian section** | 1 (4) | 1 (5) | 18 (78)[f] | 14 (58)[e] |
| **Multiple birth** | 0 | 0 | 12 (52)[h] | 8 (33)[g] |
| **APGAR score at 1 min** | 9 (5-9) | 9 (5-10) | 8 (3-10) | 5 (1-9)[i] |
| **APGAR score at 5 min** | 10 (8-10) | 10 (7-10) | 9 (3-10)[k] | 7 (3-9)[j] |
| **Installation of surfactant** | N/A | N/A | 3 (13) | 8 (33) |
| **Mechanical ventilation, days** | N/A | N/A | 0 (0-5) | 4 (0-38)[l] |
| **CPAP, days** | N/A | N/A | 3 (0-19) | 41 (3-70)[m] |
| **Supplemental $O_2$, days** | N/A | N/A | 3 (1-26) | 67 (28-180) |

Data is presented as median (min-max) or numbers (%). Abbreviations: BPD: bronchopulmonary dysplasia, N/A not applicable, CPAP= continuous positive airway pressure.

a: BPD vs healthy and asthma (p<0.001), b: preterm vs healthy and asthma (p<0.001), c: BPD vs healthy and asthma (p<0.001), d: preterm vs healthy and asthma (p<0.001), e: BPD vs healthy and asthma (p<0.001), f: preterm vs healthy and asthma (p<0.001), g: BPD vs healthy and asthma (p<0.05), h: preterm vs healthy and asthma (p<0.001), i: BPD vs healthy and asthma (p<0.001), BPD vs healthy (p<0.001) and asthma (p<0.01), k: preterm vs healthy (p<0.05), l: BPD vs preterm (p<0.001), m: BPD vs preterm (p<0.001), n: BPD vs preterm (p<0.001).

**Table E2: Abnormal lung features defined according to the nomenclature in ''Imaging of disease of the Chest", Armstrong et al, third edition.**

12 futures were included in the HRCT scoring system 1) Linear subpleural opacities 2) Triangular subpleural opacities 3) Reticular subpleural opacities 4) Local hypoatttenuation/ Mosaic perfusion 5) Air trapping 6) Emphysema 7) Bronchial wall thickening 8) Architectural distortion 9) Consolidation 10) Bronchiectasis 11) Parenchymal noduli 12) Subpleural noduli.

| | |
|---|---|
| Linear-, triangular and reticular subpleural opacities | Refers to thin lines, 1-3 mm thickness, lying less than 1 cm from the pleural surface. |
| Local hypoattenuation on inspiration/ mosaic pattern | This pattern appears as patchwork of regions of differing attenuation that may represent patchy interstitial disease, obliterative small- airways disease or occlusive vascular disease. Local hypoattenuation was defined on end inspiratory scans |
| Air trapping | Retention of air in the lung and seen on end-expiration scans. Comparison between inspiratory and expiratory CT scans can be helpful to differentiate between decreased attenuation due to hypoperfusion (mosaic pattern). |
| Emphysema | Refers to permanently enlarged airspaces distal to the terminal bronchiole with destruction of alveolar wall |
| Bronchial wall thickening | Bronchial wall thickening was considered to bepresent when both central bronchi and peripheral wall thickening were present. |
| Architectural distortion | Characterized by abnormal displacement of bronchi, vessels, fissures or septa caused by diffuse lung disease |
| Consolidation | Alveolar filling process that replaces air within the affected airspaces, increasing in pulmonary attenuation and obscuring the margins of adjacent airways and on CT scans |
| Bronchiectasis | Irreversible localized or diffuse bronchial dilatation |
| Parenchymal and subpleural noduli | Characterized as a rounded opacity, well or poorly defined, measuring up to 3 cm in diameter. Subpleural noduli was lying less than 1 cm from the pleural surface. |
| Broncho-artery ratio | Defined as the diameter of the bronchial lumen divided by the diameter of its accompanying artery. Increased broncho-arterial ratio of >1, decreased broncho arterial ratio <0,65. |

Figure E1: Bronchoarterial ratio in subjects with BPD compared to healthy, asthma and preterm subjects

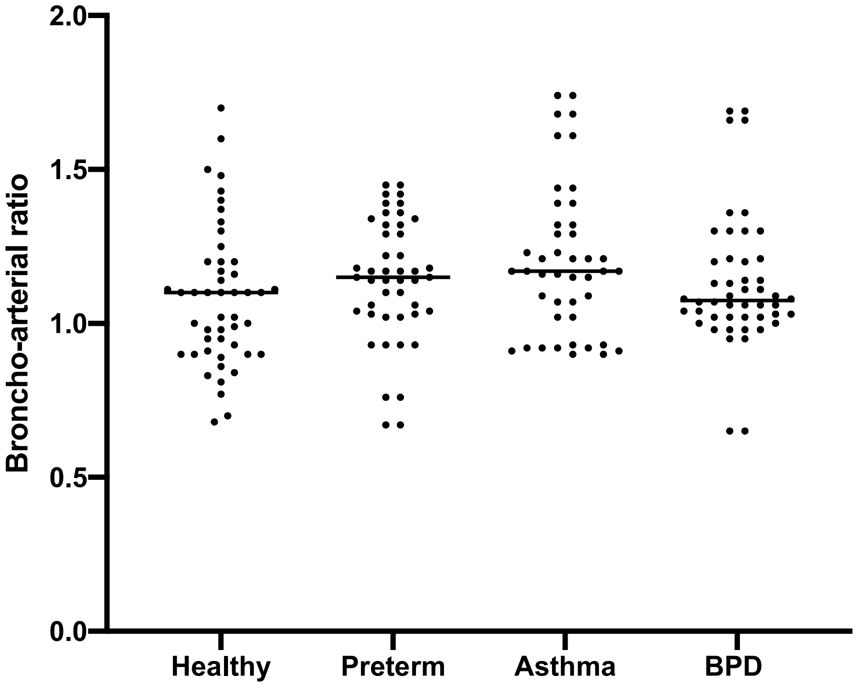


Figure E2: Correlation between lung volume on CT and diffusion capacity (DLCOadj).

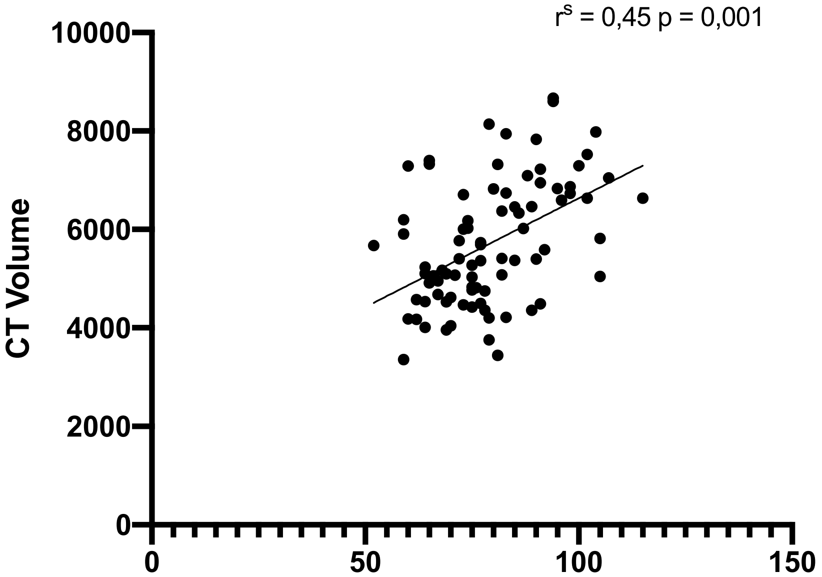

Figure E3. Correlation between mean Hounsfield unit (mean HU) and adjusted diffusion capacity (DLCOadj) in BPD subjects.

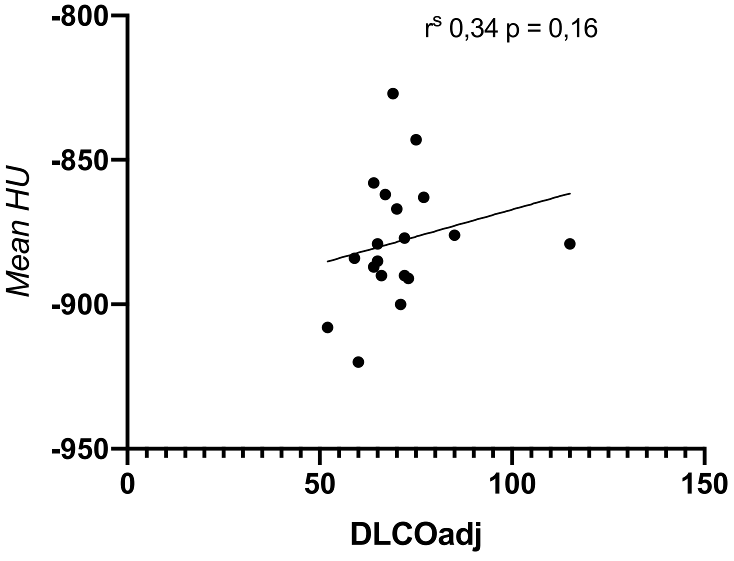
